# ThaparUni at #SMM4H 2023: Synergistic Ensemble of RoBERTa, XLNet, and ERNIE 2.0 for Enhanced Textual Analysis^1^

**DOI:** 10.1101/2023.11.10.23298362

**Authors:** Sharandeep Singh, Jatin Bedi

**Affiliations:** Thapar Institute of Engineering and Technology, Patiala, Punjab, India

## Abstract

This paper presents the system developed by Team ThaparUni for the Social Media Mining for Health Applications (SMM4H) 2023 Shared Task 4. The task involved binary classification of English Reddit posts, focusing on self-reporting social anxiety disorder (SAD) diagnoses. The final system employed a combination of three models: RoBERTa, ERNIE, and XLNet, and results obtained from all three models were integrated. The results, specifically in the context of mental health-related content analysis on social media platforms, show the possibility and viability of using multiple models in binary classification tasks.

## Introduction

Social media platforms have revolutionized the way individuals communicate, providing an invaluable window into their thoughts, feelings, and experiences. As it presents a rare opportunity to obtain insight into individuals’ viewpoints on their health situations, this abundant source of data has inspired growing interest in the field of healthcare. One such area of interest is Social Anxiety Disorder (SAD), a prevalent mental health condition with substantial implications for patients’ well-being and quality of life.

## Data

The dataset used for this study is obtained from the r/socialanxiety subreddit, a community where individuals share their thoughts, feelings, and struggles related to Social Anxiety Disorder. The dataset encompasses a total of 6390 posts authored by users aged between 12 to 25 years, each accompanied by a unique identifier, textual content, and a diagnosis label.

## Data Preprocessing

The data obtained in this case was noisy, containing errors such as typos, grammatical errors, and slang. This noise made it difficult to learn and understand the correct patterns. Therefore, the data was pre-processed, following the process, (i) Conversion of all characters in the text to lowercase, (ii) All URLs (starting with “https://”) were replaced with a single space, (iii) All punctuation marks were removed from the text, (iv) All non-word characters, except whitespace and periods with spaces, were replaced, effectively filtering out special characters and retaining only meaningful words, (v) Words containing numbers were removed. In the dataset, there were two types of textual content: posts and comments.

While posts have a title and text associated with them, comments only have the text without any separate title. Since both posts and comments contain valuable information for analysis, a consistent way to process and analyze the textual content from both data types was needed. To achieve this, a new column called ‘title_text’ was created by combining the existing ‘title’ and ‘text’ columns for posts. Only the ‘text’ column was considered for comments since comments do not have separate titles. By combining the ‘title’ and ‘text’ columns for posts, it was ensured that the textual content from both posts and comments was integrated into a single column, making the subsequent data processing steps consistent for both types of content.

## Models

The BERT models used in this approach were all pre-trained. This pre-training helped the models to learn the relevant features of language, which were then used for the text classification task. The use of a combination of three models was employed in this case to leverage their individual strengths and enhance the overall performance of the system. They have been shown to perform well on a variety of natural language processing tasks, including text classification. RoBERTa is known for its ability to learn long-range dependencies between words. XLNet is known for its ability to handle noisy and ambiguous text. ERNIE 2.0 is known for its ability to understand the context of words and phrases. The ensemble technique involves training multiple neural networks for the same task. In this approach, RoBERTa, ERNIE 2.0, and XLNet models were individually fine-tuned, and their predicted labels for each sample were extracted. A voting system that uses a hard voting scheme was then applied to these labels, selecting the one that occurred most frequently as the final label for each sentence, as shown in Figure 1.

**Figure 1.**
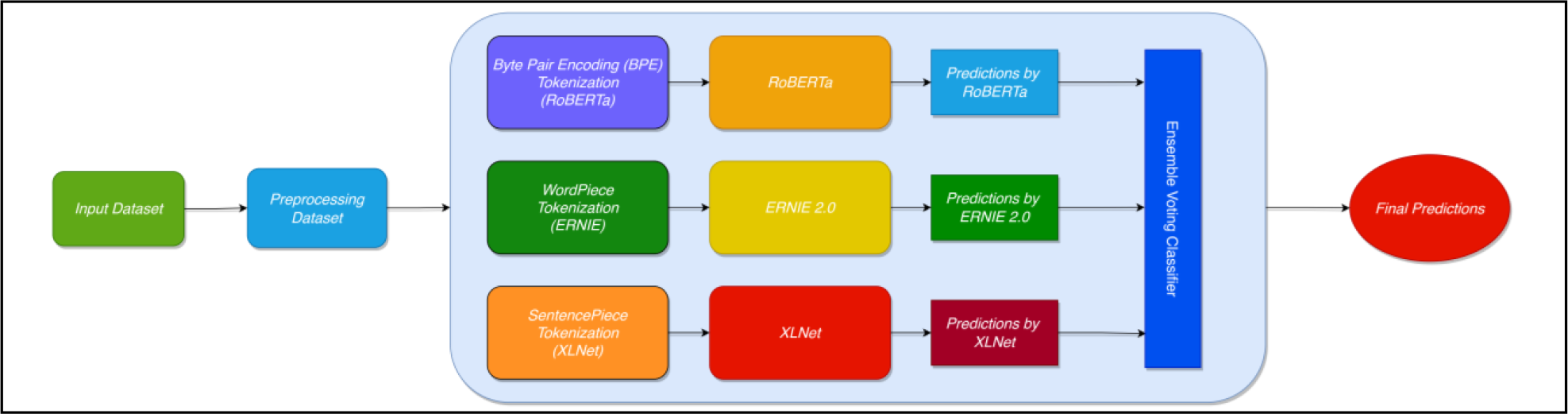
The Architecture of Ensemble Voting Classifier.

Pretrained models from HuggingFace^7^ were adopted and fine-tuned using PyTorch^8^. Each of the models, RoBERTa, ERNIE, and XLNet, was trained for 5 epochs with the learning rate decay of 0.01 using AdamW optimizer^6^ and maximum sequence size set to 512. The training process was performed on a Google Colab notebook with a T4 GPU, with 16 GB of memory and 48 CUDA cores.

## Results

Table 2 exhibits the results achieved by the best-performing systems and the mean and median performance on the test set across all teams’ submissions for Task 4. The system showcased commendable performance, outperforming the test set median F1 score by 1.8% and the mean F1 score by 4.9%. These evaluations provided valuable insights into the system’s strengths and limitations, guiding towards refining, and optimizing its performance for future applications. It is worth noting that although RoBERTa’s performance on the test set is about the same as the ensemble model, but the ensemble model was better at identifying posts that actually self-report a social anxiety disorder diagnosis, considering the higher recall.

**Table 1:** Overview of Task 4 Data.

| Dataset | Total Number of Samples | Number of Positive Samples | % of Positive Samples |
| --- | --- | --- | --- |
| Train | 4500 | 1702 | 37.8% |
| Validation | 640 | 330 | 36.7% |
| Test | 1250 | - | - |

**Table 2.**
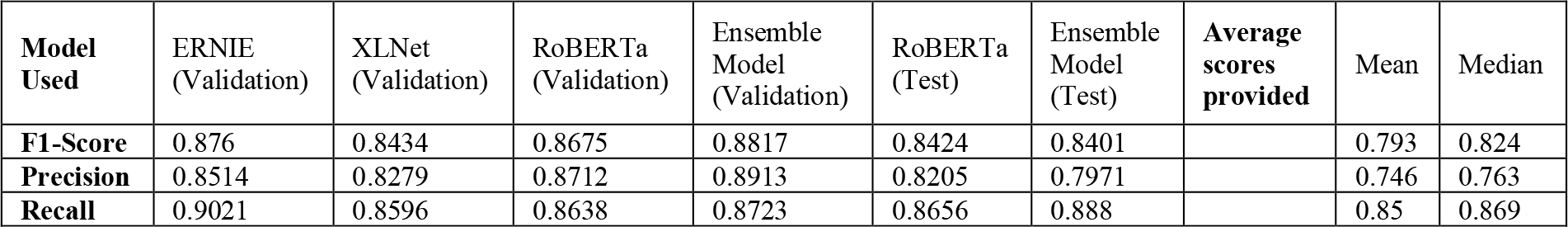
Binary Classification results on Task 4 (F1-Score, Precision, and Recall)

| Model Used | ERNIE (Validation) | XLNet (Validation) | RoBERTa (Validation) | Ensemble Model (Validation) | RoBERTa (Test) | Ensemble Model (Test) | Average scores provided | Mean | Median |
| --- | --- | --- | --- | --- | --- | --- | --- | --- | --- |
| **F1-Score** | 0.876 | 0.8434 | 0.8675 | 0.8817 | 0.8424 | 0.8401 |  | 0.793 | 0.824 |
| **Precision** | 0.8514 | 0.8279 | 0.8712 | 0.8913 | 0.8205 | 0.7971 |  | 0.746 | 0.763 |
| **Recall** | 0.9021 | 0.8596 | 0.8638 | 0.8723 | 0.8656 | 0.888 |  | 0.85 | 0.869 |

## Conclusion

In conclusion, the participation of Team ThaparUni, in the SMM4H Tasks showcased the success of the ensemble model, employing a strategic combination of cutting-edge RoBERTa, ERNIE 2.0, XLNet, and fine-tuned language models. This novel approach yielded exceptional results, evident from the commendable 84.2% F1 Score achieved in Task 4. By leveraging the strengths of various models, the significance of ensemble techniques in enhancing the performance of natural language processing tasks was demonstrated, particularly in the realm of social media mining for health applications. The remarkable efficacy of the approach underscores the potential of interpretable machine learning techniques in understanding patient perspectives, disease burden, and unmet medical needs through social media analysis.

## Data Availability

All data produced in the present study are available upon reasonable request to the organizers of the SMM4H Workshop.

## Footnotes

1 This paper has been peer-reviewed and accepted for presentation at the #SMM4H 2023 Workshop.

## References

1. Liu Y, Ott M, Goyal N, Du J, Joshi M, Chen D, Levy O, Lewis M, Zettlemoyer L, Stoyanov V. RoBERTa: A robustly optimized BERT pretraining approach. arXiv preprint arXiv:1907.11692. 2019 Jul 26.

2. Sun Y, Wang S, Li Y, Feng S, Tian H, Wu H, Wang H. ERNIE 2.0: A continual pre-training framework for language understanding. InProceedings of the AAAI conference on artificial intelligence 2020 Apr 3 (Vol. 34, No. 05, pp. 8968–8975).

3. Zhang Z, Han X, Liu Z, Jiang X, Sun M, Liu Q. ERNIE: Enhanced language representation with informative entities. arXiv preprint arXiv:1905.07129. 2019 May 17.

4. Klein AZ, Banda JM, Guo Y, Flores Amaro JI, Rodriguez-Esteban R, Sarker A, Schmidt AL, Xu D, Gonzalez-Hernandez G. Overview of the eighth Social Media Mining for Health Applications (#SMM4H) Shared Tasks at the AMIA 2023 Annual Symposium. In: Proceedings of the Eighth Social Media Mining for Health Applications (#SMM4H) Workshop and Shared Task; 2023.

5. Yang Z, Dai Z, Yang Y, Carbonell J, Salakhutdinov RR, L. QV. XLNet: Generalized autoregressive pretraining for language understanding. Advances in neural information processing systems. 2019;32.

6. Loshchilov I, Hutter F. Decoupled weight decay regularization. arXiv preprint arXiv:1711.05101. 2017 Nov 14.

7. Wolf T, Debut L, Sanh V, Chaumond J, Delangue C, Moi A, Cistac P, Rault T, Louf R, Funtowicz M, Davison J. Huggingface’s transformers: State-of-the-art natural language processing. arXiv preprint arXiv:1910.03771. 2019 Oct 9.

8. Paszke A, Gross S, Massa F, Lerer A, Bradbury J, Chanan G, Killeen T, Lin Z, Gimelshein N, Antiga L, Desmaison Pytorch: An imperative style, high-performance deep learning library. Advances in neural information processing systems. 2019;32.

